# An International Longitudinal Natural History Study of Danon Disease Patients: Unique Cardiac Trajectories Identified Based on Sex and Heart Failure Outcomes

**DOI:** 10.1101/2024.05.21.24307720

**Authors:** Kimberly N. Hong, Emily Eshraghian, Tarek Khedro, Alessia Argirò, Jennifer Attias, Garrett Storm, Melina Tsotras, Tanner Bloks, Isaiah Jackson, Elijah Ahmad, Sharon Graw, Luisa Mestroni, Quan M. Bui, Jonathan Schwartz, Stuart Turner, Eric D. Adler, Matthew Taylor

**Affiliations:** University of California, San Diego, La Jolla, California; University of Minnesota Medical School, Minneapolis, Minnesota; Royal College of Surgeons in Ireland, Dublin, Ireland; Cardiomyopathy Unit, University of Florence, Florence, Italy; University of Colorado Anschutz Medical Center, Denver, Colorado; Rocket Pharmaceuticals Inc., New York, New York

## Abstract

**Introduction:** Danon disease (DD) is a rare X-linked dominant cardioskeletal myopathy caused by mutations in the lysosome-associated membrane protein-2 gene. Though the severe morbidity of disease in males is well established, longitudinal studies describing the trajectory of cardiovascular disease in both sexes have not been performed. Herein we performed an analysis using the International Danon Disease Registry, a retrospective dataset that includes longitudinal data from a cohort of males and females with DD.

**Methods:** Data were from the International Danon Disease Registry and includes patients from 2005 to 2022. Records were obtained from the first episode of care to the date of enrollment and included demographics, clinical characteristics, echocardiographic and laboratory values, and outcomes. The primary outcome in this study was a heart failure (HF) composite defined as either transplant (TXP), left ventricular assist device (LVAD) or death.

**Results:** The analysis included 116 DD patients: female (n=64, 55%) and male (n=52, 45%). Median age of diagnosis for the entire cohort was 15.2 years (10.0-25.2 years), and 21.9 years (15.0-34.9 years) and 12.4 years (7.1-15.6 years) for females and males respectively. The incidence of HF outcome was higher in males compared to females (p<0.001). Regarding trends in echocardiographic parameters over time, LVEF decreased and LVEDD increased regardless of sex and HF outcome, however, rate of change was increased in patients who experienced a HF outcome in both sexes. Increasing LV mass occurs in males but not females. Consistent with this, LV wall hypertrophy continues in males who have not yet experienced a HF outcome and stabilizes prior to HF outcome, while females have progressive LV thinning regardless of HF outcome. Analyses stratified by age of HF outcome in females found two distinct groups of females, one who experienced HF outcome prior to 26 years of age and another after.

**Conclusions:** In this largest longitudinal natural history study of DD to date, we confirmed that males present on average a decade earlier and demonstrate more progressive cardiac hypertrophy and heart failure than females. Of note, there may be a subset of females who are phenotypically similar to males with profound LV hypertrophy that appears to stabilize or regress prior to HF outcome. Correlations between structural changes including LV hypertrophy, dilation and dysfunction and disease progression may allow for risk stratification of Danon patients and refinement of treatment algorithms while also informing therapeutic trial design.

## INTRODUCTION

Danon disease (DD) is a rare X-linked dominant cardioskeletal myopathy caused by mutations in the lysosome-associated membrane protein-2 (*LAMP2*) gene.^1^ It is characterized by a severe cardiomyopathy associated with skeletal myopathy and cognitive impairment. DD in males is typically lethal without cardiac transplantation, and while disease severity is variable in females, there is a subset of females who require cardiac transplantation at similar ages to males.^2–5^ As an X-linked disorder, disease course varies by sex, with males typically developing extracardiac symptoms early in childhood. Cardiac manifestations present soon after and reach end-stage in males by the second or third decade of life. Female extracardiac symptoms are variable, with cardiac manifestations in the 2^nd^ or 3^rd^ decade of life typically being the first symptoms.^5–8^ Understanding natural disease progression is critical to improving outcomes in DD patients, as it provides not only a basis for assessing the efficacy of treatments, but also allows appropriate identification of patients for evolving therapeutics. The purpose of this paper is to describe differences in disease progression as characterized by longitudinal changes in echocardiographic and laboratory parameters prior to death, heart transplantation or ventricular assist device in males and females with DD.

## METHODS

Data were from the International Danon Disease Registry that includes patients from the United States and Europe consented between 2005 and 2022. There were two enrolling sites in the United States, University of Colorado and University of California, San Diego. Patients were referred by their physician or self-referred through patient community and outreach sites including www.DanonDisease.org, https://rarediseases.org/rare-diseases/danon-disease/, and two Facebook groups (Danon Disease Support Group and Danon Disease Support). All patients were required to have a history of positive genetic testing (defined as pathogenic or likely pathogenic *LAMP2* variations) or confirmed genealogy to a known DD carrier prior to enrollment. Specific genetic data was collected for 50% (n=60) of the patient cohort. Records were obtained from the first episode of care to the date of enrollment. These data include patients born between the years of 1943 and 2020.

Data collected included demographics, clinical characteristics such as involvement of extracardiac systems and concomitant cardiac conditions, echocardiographic parameters, laboratory values, cardiovascular implantable electronic device (CIED) implantation and ablation. Specific echocardiographic parameters that were collected included left ventricular (LV) thickest wall which was defined as the largest measurement between septal and posterior wall thickness, LV ejection fraction (LVEF), LV end diastolic diameter (LVEDD), which is the distance measured between the LV septal and posterior walls in the parasternal view, and LV mass which was calculated using the Cube formula (LV mass=(0.8*1.04[(IVS+LVEDD+PWd)^3-LVEDD^3]+0.6 grams), wherein LV measurements are in cm).^9^ Outcome measures include heart transplantation, left ventricular assist device and death. Z-scores were calculated for LV septal and posterior wall, LVEDD and LV mass measurements in patients with echocardiograms completed prior to the age of 18 years.

Laboratory values, including skeletal muscle, cardiac, and liver biomarkers, were collected. These included creatine phosphokinase (mcg/L), brain natriuretic peptide (pg/mL), aspartate transaminase (AST, IU/L), alanine aminotransferase (ALT, IU/L), alkaline phosphatase (IU/L) and total bilirubin (mg/dL).

All data were analyzed with the statistical software package Stata 17 (Stata Corp, College Station, TX). Continuous variables were reported as either mean with standard deviation or median with interquartile range depending on normality testing and compared with either Student’s t-test or Wilcoxon Rank Sum non-parametric test as appropriate. To compare categorical variables, the Pearson’s *X*^2^ was used. The conventional probability value of 0.05 or less was used to determine statistical significance. All reported *P-*values are two-sided. For longitudinal analysis, Wilcoxon Rank Sum testing was used to compare first and last echocardiograms and linear mixed models were used to test for trends in echocardiography parameters over time/age. For the echocardiography measures, separate mixed models were run for measures with pediatric z-scores (included echocardiograms completed prior to the age of 18 years of age) and for all values (included all echocardiograms). Age was the fixed effect, and the individual patient the random effect. For survival analysis, Kaplan-Meier analysis was used for time-to-HF event analysis, with log-rank test used to assess differences across sex. Patients alive at last follow-up were censored on the day of last known follow-up.

## RESULTS

The analysis included 116 DD patients and was stratified by sex: female (n=64, 55%) and male (n=52, 45%). [Table 1] Median age of diagnosis for the entire cohort was 15.2 years (10.0-25.2 years), and 21.9 years (15.0-34.9 years) and 12.4 years (7.1-15.6 years) for females and males respectively. Median age at last follow-up time for the entire cohort was 23.0 years (IQR: 16.5 – 34.5 years), and 32.0 years (IQR: 22.5 – 46.0 years) and 18.5 years (14.0 – 23.0 years) for females and males respectively. Median follow up time was 5.82 years (2.67 – 11.76 years) for the entire cohort, and 5.82 years (2.67 – 14.96 years) and 5.86 years (2.93 – 8.51 years) for females and males respectively. The majority of patients in the registry were White (72.4%). Regarding arrhythmia characteristics, 19.8% (n=23) of patients had WPW, 18.1% (n=21) had atrial fibrillation (AF) and 38.8% (n=45) of patients had a CIED; there were no differences in prevalence of WPW (p=0.429) or CIED (p=0.653) implantations between males and females. AF occurred more frequently in females compared to males (p=0.004). Within patients who received a CIED, 2 males (3.8%) were implanted with pacemakers alone; the remaining patients were implanted with an AICD, of which 5.2% (n=2) were for secondary prevention.

**Table 1.** Summary statistics for patients stratified by sex.

|  | Males |  | Females |  | Total |  |
| --- | --- | --- | --- | --- | --- | --- |
|  | # | IQR/% | # | IQR/% | # | IQR/% |
| n | 52 | 44.8 | 64 | 55.2 | 116 |  |
| Age (median) | 19 | 16-23 | 26 | 14-39 | 20 | 16-27 |
| Hispanic | 3 | 5.8 | 1 | 1.6 | 4 | 3.4 |
| White | 38 | 73.1 | 46 | 71.9 | 84 | 72.4 |
| Black | -- | -- | -- | -- | -- | -- |
| Asian | 1 | 1.9 | 3 | 4.7 | 4 | 3.4 |
| WPW | 12 | 23.1 | 11 | 17.2 | 23 | 19.8 |
| LVEF (%) |  |  |  |  |  |  |
| At first echo | 66.1 | 60-72 | 60.6 | 51.5-72.5 | 62.2 | 55-72 |
| At last echo | 57.5 | 42.7-68 | 55 | 38.7-64.8 | 55 | 38.7-65 |
| LVWT (cm) |  |  |  |  |  |  |
| At first echo | 1.11 | (0.86-1.93) | 1.1 | (0.81-1.50) | 1.1 | (0.83-1.60) |
| At last echo | 1.36 | (0.95-2.18) | 1.19 | (1.00-1.73) | 1.29 | (1.00-1.85) |
| LV Mass (g) |  |  |  |  |  |  |
| At first echo | 140 | (80-263) | 159 | (97-242) | 91 | (148-242) |
| At last echo | 430 | (120-647) | 235 | (140-287) | 238 | (140-430) |
| HF Outcome | 21 | 40.4 | 14 | 21.9 | 35 | 30.2 |
| TXP | 17 | 32.7 | 13 | 20.3 | 30 | 25.9 |
| LVAD | 2 | 3.8 | 2 | 3.1 | 4 | 3.4 |
| Death | 7 | 13.5 | 2 | 3.1 | 6 | 5.2 |
| Age at HF Outcome | 19 | (16-23) | 26 | (14-39) | 20 | (16-27) |
| CIED | 19 | 36.5 | 26 | 40.6 | 45 | 38.8 |
| Pacemaker | 2 | 3.8 | 0 | 0.0 | 2 | 1.7 |
| AICD | 17 | 32.7 | 26 | 40.6 | 43 | 37.1 |
| 2° prevention | 2 | 3.8 | 4 | 6.3 | 6 | 5.2 |
| AICD: automatic implantable cardioverter defibrillator, CIED: Cardiac Implantable Electrical Devices, IQR: interquartile range, LVEF: left ventricular ejection fraction, LVAD: left ventricular assist device, TXP: heart transplant, WPW: Wolff-Parkinson-White |  |  |  |  |  |  |

### Heart Failure Outcomes

The incidence of the HF outcome was higher in males compared to females (p=0.031), and actuarial median age of HF outcome was younger in males compared to females with median time to HF outcome being 25 years in males and 63 years in females (P<0.001). [Figure 1] While age of HF outcome in males (19 years (16-23 years)) was numerically lower compared to females (26 years (14-39 years)) who did experience HF outcome, this did not reach statistical significance (p=0.162). This may be due to a possible bimodal distribution in age at which females experience a HF outcome [Figure 2]. Median age of HF outcome in females with HF outcome ≧ 26yo (n=7) and <26yo (n=7) was 39 (28-47yrs) and 14 (11-20yrs) respectively (p=0.002). Additional analyses stratifying females by age of HF outcome are presented below.

**Figure 1.**
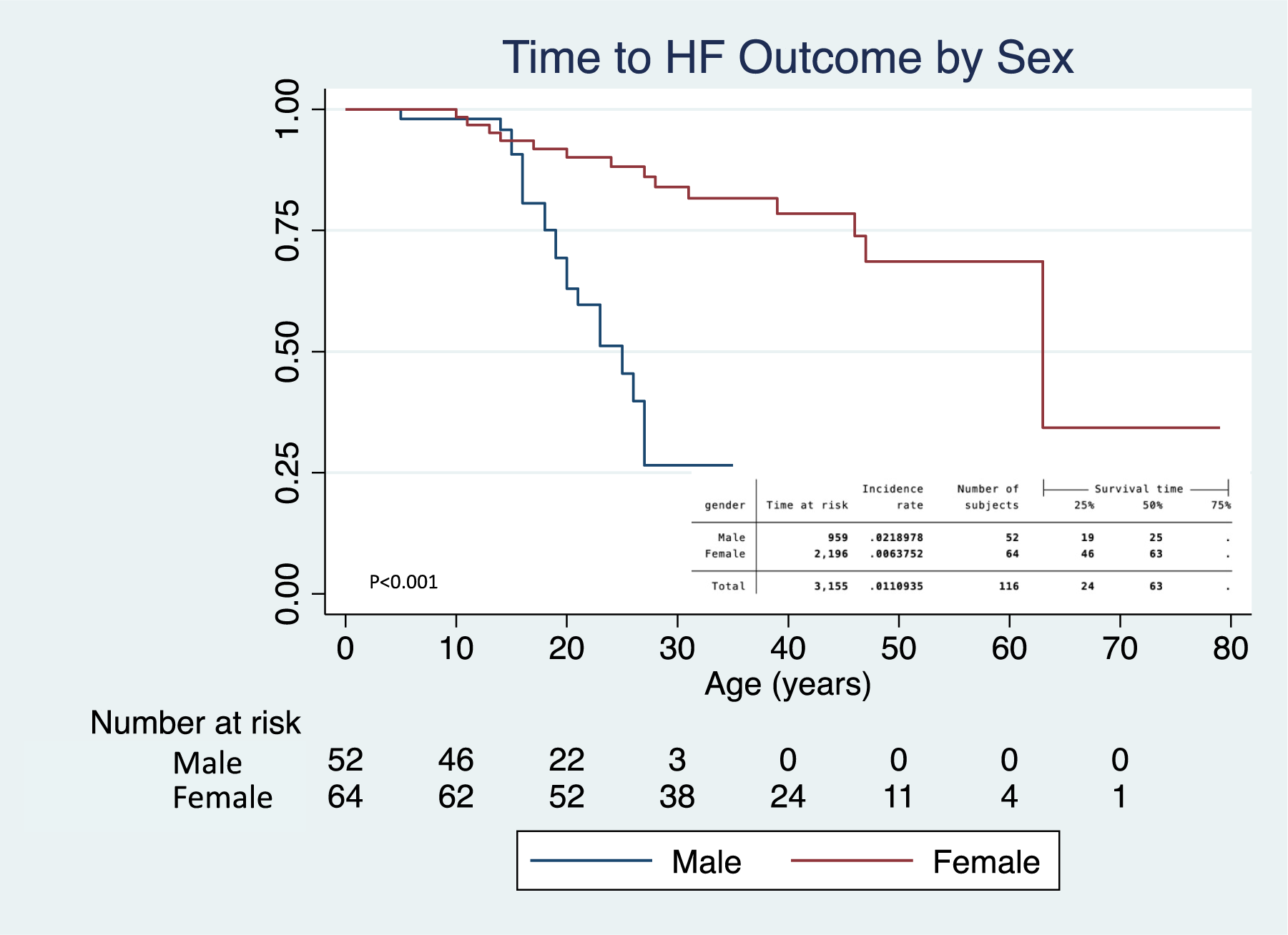
Kaplan Meier Time to HF Outcome Analysis Stratified by Sex

**Figure 2.**
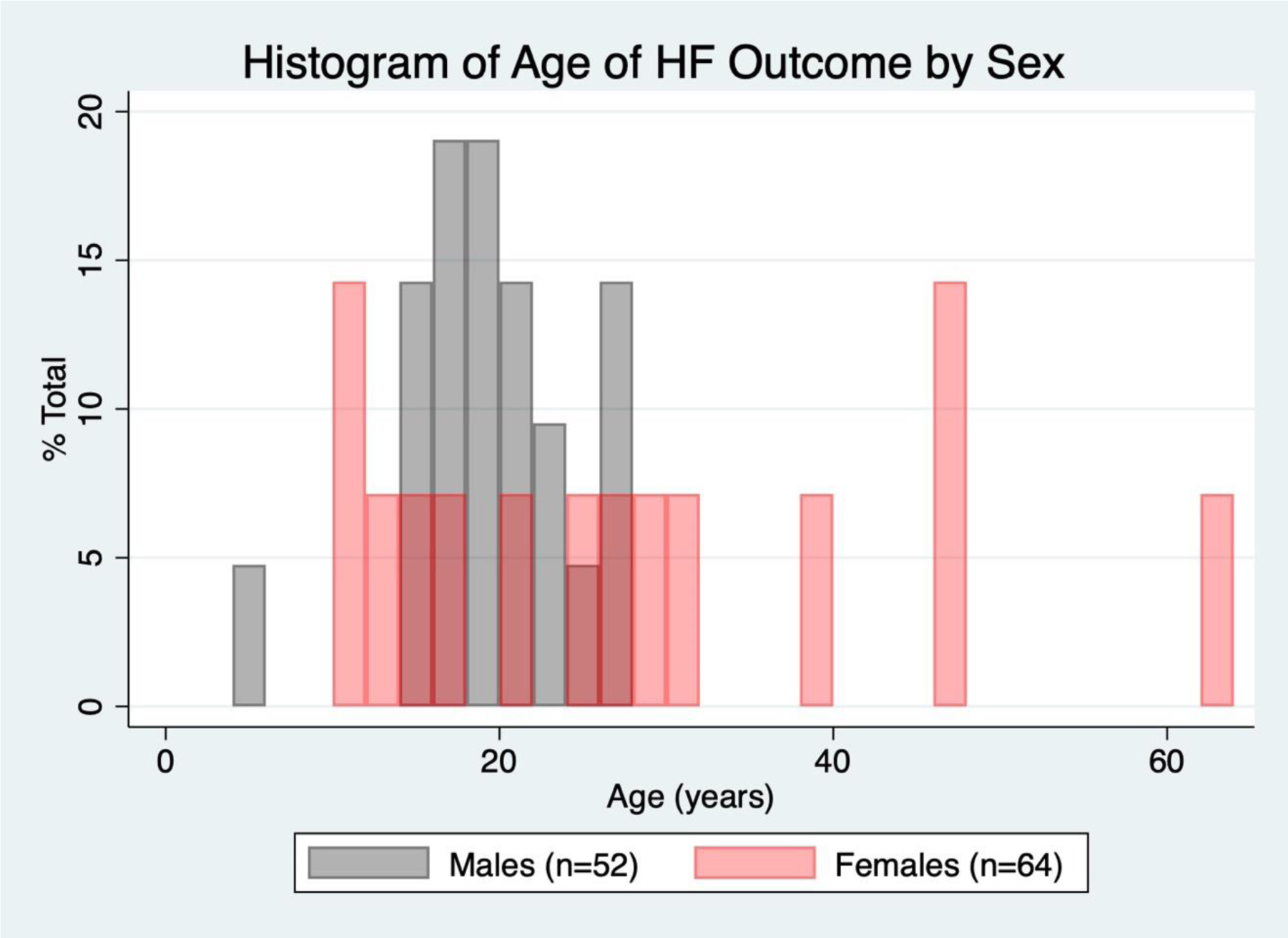
Histogram of age at heart failure outcome stratified by sex. There is a suggestion of a bimodal distribution for HF outcome in females.

### Echocardiographic Parameter Analyses

A total of 95 (82%) patients had echocardiographic results, of which 75 (92%) had serial echocardiograms. Ages of first echocardiogram in patients with serial echocardiograms were 15.9 years (11.3-19.9 years) and 25.8 years (17.0-42.2 years) in males and females respectively. Median time from first to last echocardiogram in all patients was 4.7 years (IQR: 1.1-8.9 years), and 4.2 years (IQR: 0.7-6.1 years) and 6.1 years (IQR 1.2-11.3 years) in males and females respectively.

#### LV Function

In longitudinal analyses of LVEF which compared first and last LVEF by echocardiogram, LVEF decreased in both males (p=0.003) and females (p=0.02) over time. [Table 2] In analyses stratified by HF outcome, based on comparison of coefficients and 95% confidence intervals from the linear mixed effects models, the rate of LVEF decline is higher in patients who experience a HF outcome compared to those who have not. [Figure 3] Specifically, mixed effects model coefficients in both males and females, suggest a decrease in LVEF by ∼2.4% per year in patients that experience a HF outcome. [Table 3] Notably, in patients who experienced a HF outcome, LVEF from baseline to last echocardiogram decreased from 58% to 33% (p=0.021) and from 57% to 37% (p=0.054) in males and females respectively. [Table 2]

**Figure 3.**
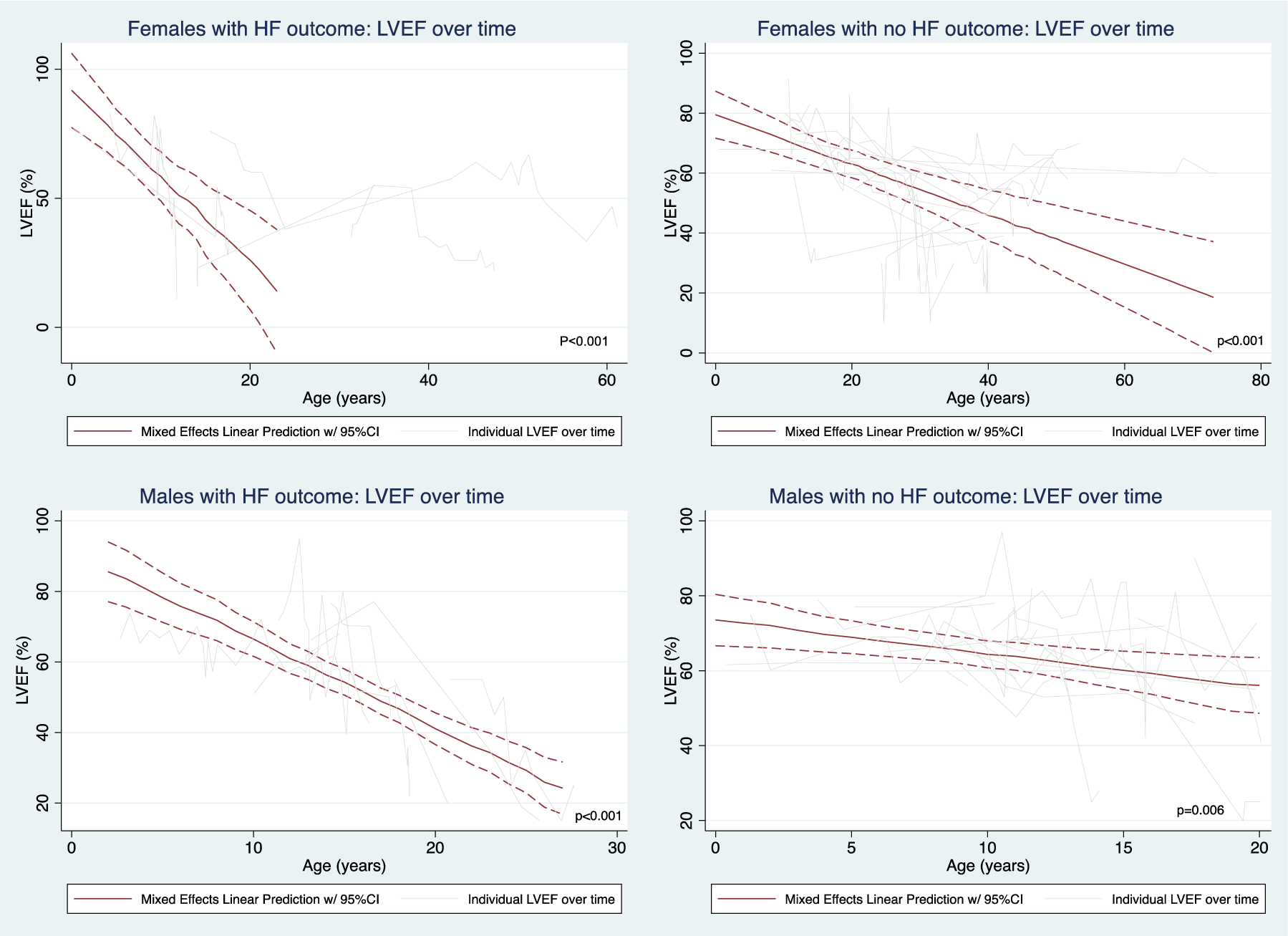
LVEF changes over time, as represented by linear mixed effects regression plot and individual spaghetti plots, stratified by sex and HF outcome. In females experiencing HF outcome, mixed effects regression plot for patients experiencing HF outcome prior to the age of 26 years old was graphed.

**Table 2.** Echocardiographic measurements at first and last exam stratified by sex. Within and between sex comparisons between first and last measurement were completed and p-values reported. Statistically significant p-values were bolded.

|  | Total |  |  | HF Outcome (+) |  |  | HF Outcome (-) |  |  |  |
| --- | --- | --- | --- | --- | --- | --- | --- | --- | --- | --- |
|  | Median | IQR | p-value* | Median | IQR | p-value* | Median | IQR | p-value* | p-value^ |
| Males |  |  |  |  |  |  |  |  |  |  |
| LVEF (%) |  |  |  |  |  |  |  |  |  |  |
| First | 66 | (57-71) | **0.003** | 58 | (50-68) | **0.021** | 67 | (62-76) | **0.0135** | **0.030** |
| Last | 55 | (28-64) |  | 33 | (20-52) |  | 58 | (50-68) |  | **0.011** |
| Thickest LV wall (cm) |  |  |  |  |  |  |  |  |  |  |
| First | 1.2 | (0.9-1.9) | 0.245 | 1.7 | (1.2-2.4) | 0.713 | 1.1 | (0.8-1.5) | 0.173 | **0.015** |
| Last | 1.5 | (1.0-2.5) |  | 1.9 | (1.6-2.5) |  | 1.4 | (1.0-2.3) |  | 0.268 |
| LV Mass (g) |  |  |  |  |  |  |  |  |  |  |
| First | 154 | (87-413) | 0.12 | 327 | (209-541) | 0.285 | 111 | (80-163) | 0.172 | **0.005** |
| Last | 275 | (143-613) |  | 596 | (285-690) |  | 161 | (77-377) |  | **0.020** |
| LVEDD (cm) |  |  |  |  |  |  |  |  |  |  |
| First | 3.9 | (3.4-4.6) | 0.185 | 4 | (3.6-4.6) | 0.115 | 3.8 | (3.4-4.7) | 0.712 | 0.851 |
| Last | 4.4 | (3.7-5.3) |  | 6.3 | (4.8-6.6) |  | 4 | (3.7-4.7) |  | **0.038** |
| Females |  |  |  |  |  |  |  |  |  |  |
| LVEF (%) |  |  |  |  |  |  |  |  |  |  |
| First | 61 | (55-70) | **0.020** | 57 | (52-77) | 0.054 | 61 | (55-69) | 0.221 | 0.929 |
| Last | 56 | (35-65) |  | 37 | (23-47) |  | 56 | (47-66) |  | **0.014** |
| Thickest LV wall |  |  |  |  |  | 0.962 |  |  | 0.105 |  |
| First | 1.1 | (0.8-1.5) | 0.162 | 1.1 | (1.0-1.6) |  | 1.1 | (0.8-1.4) |  | 0.416 |
| Last | 1.2 | (1.0-1.7) |  | 1.2 | (0.9-2.0) |  | 1.2 | (1.0-1.7) |  | 0.937 |
| LV Mass (g) |  |  |  |  |  |  |  |  |  |  |
| First | 165 | (97-262) | 0.132 | 296 | (80-320) | 0.723 | 157 | (106-227) | 0.088 | 0.381 |
| Last | 224 | (140-272) |  | 232 | (218-288) |  | 187 | (132-272) |  | 0.554 |
| LVEDD |  |  |  |  |  |  |  |  |  |  |
| First | 4.3 | (3.6-4.9) | **0.012** | 4.4 | (2.8-5.2) | **0.018** | 4.3 | (3.9-4.8) | 0.144 | 0.974 |
| Last | 4.7 | (4.1-5.8) |  | 5.9 | (5.5-6.4) |  | 4.5 | (4.0-5.3) |  | **0.003** |
| *comparing first and last echocardiograms; ^comparing HF Outcome (+) and HF Outcome (-)<br>LVEF: left ventricular ejection fraction, LVEDD: left ventricular end diastolic dimension |  |  |  |  |  |  |  |  |  |  |

**Table 3.**
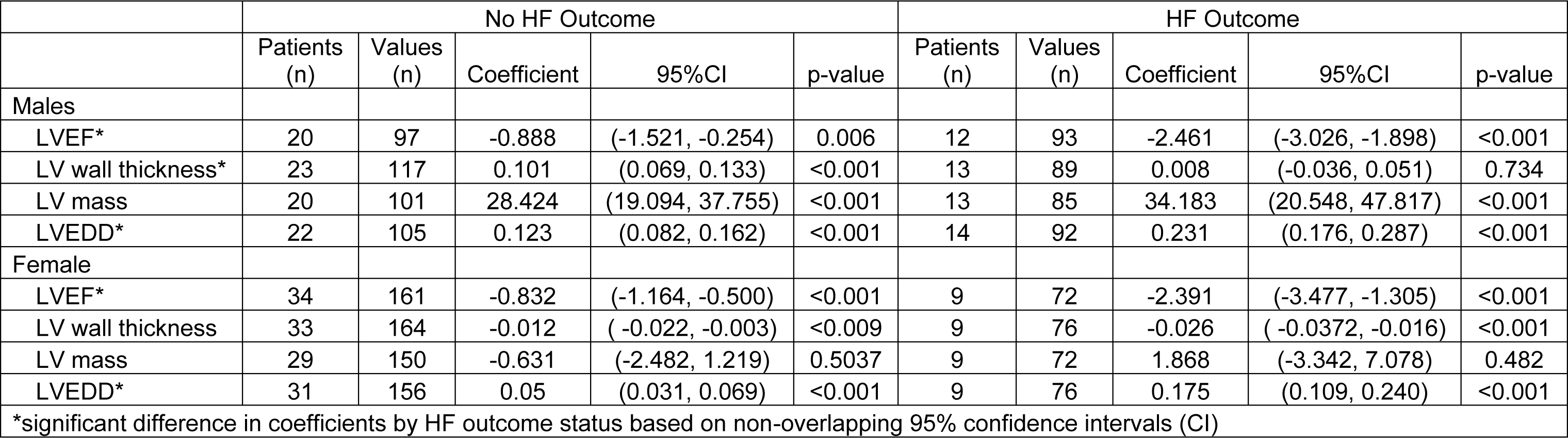
Longitudinal trend ana lysis of echocardiographic parameters over time. Coefficients derived from mixed effect regression models represent the magnitude of change in the parameter as patient ages.

|  | No HF Outcome |  |  |  |  | HF Outcome |  |  |  |  |
| --- | --- | --- | --- | --- | --- | --- | --- | --- | --- | --- |
|  | Patients<br>(n) | Values<br>(n) | Coefficient | 95%CI | p-value | Patients<br>(n) | Values<br>(n) | Coefficient | 95%CI | p-value |
| Males |  |  |  |  |  |  |  |  |  |  |
| LVEF* | 20 | 97 | -0.888 | (-1.521, -0.254) | 0.006 | 12 | 93 | -2.461 | (-3.026, -1.898) | <0.001 |
| LV wall thickness* | 23 | 117 | 0.101 | (0.069, 0.133) | <0.001 | 13 | 89 | 0.008 | (-0.036, 0.051) | 0.734 |
| LV mass | 20 | 101 | 28.424 | (19.094, 37.755) | <0.001 | 13 | 85 | 34.183 | (20.548, 47.817) | <0.001 |
| LVEDD* | 22 | 105 | 0.123 | (0.082, 0.162) | <0.001 | 14 | 92 | 0.231 | (0.176, 0.287) | <0.001 |
| Female |  |  |  |  |  |  |  |  |  |  |
| LVEF* | 34 | 161 | -0.832 | (-1.164, -0.500) | <0.001 | 9 | 72 | -2.391 | (-3.477, -1.305) | <0.001 |
| LV wall thickness | 33 | 164 | -0.012 | ( -0.022, -0.003) | <0.009 | 9 | 76 | -0.026 | ( -0.0372, -0.016) | <0.001 |
| LV mass | 29 | 150 | -0.631 | (-2.482, 1.219) | 0.5037 | 9 | 72 | 1.868 | (-3.342, 7.078) | 0.482 |
| LVEDD* | 31 | 156 | 0.05 | (0.031, 0.069) | <0.001 | 9 | 76 | 0.175 | (0.109, 0.240) | <0.001 |
| *significant difference in coefficients by HF outcome status based on non-overlapping 95% confidence intervals (CI) |  |  |  |  |  |  |  |  |  |  |

**Table 4.** Laboratory measurements at first and last exam stratified by sex. Within and between sex comparisons between st and last measurement were completed and p-values reported. Statistically significant p-values were bolded.

|  | Males |  |  |  | Females |  |  |  | p-value^ |
| --- | --- | --- | --- | --- | --- | --- | --- | --- | --- |
|  | n | Median | IQR/% | p-value* | n | Median | IQR/% | p-value* |  |
| CPK | 27 |  |  |  | 20 |  |  |  |  |
| At first lab draw |  | 891 | 609-1258 | 0.6 |  | 73.5 | 53-124 | 0.74 | **<0.001** |
| At last lab draw |  | 834 | 541-1868 |  |  | 77 | 42-114 |  | **<0.001** |
| BNP | 32 |  |  |  | 32 |  |  |  |  |
| At first lab draw |  | 227 | 30-999 | 0.119 |  | 383 | 141-997 | 0.726 | 0.29 |
| At last lab draw |  | 746 | 129-1208 |  |  | 353 | 200-1321 |  | 0.518 |
| AST | 23 |  |  |  | 20 |  |  |  |  |
| At first lab draw |  | 269 | 186-383 | 0.468 |  | 43 | 25-71 | 0.636 | **<0.001** |
| At last lab draw |  | 234 | 128-396 |  |  | 39 | 27-55 |  | **<0.001** |
| ALT | 32 |  |  |  | 32 |  |  |  |  |
| At first lab draw |  | 234 | 113-381 | 0.6011 |  | 28 | 22-36 | 0.696 | **<0.001** |
| At last lab draw |  | 205 | 106-388 |  |  | 26 | 21-35 |  | **<0.001** |
| Alk Phos | 32 |  |  |  | 32 |  |  |  |  |
| At first lab draw |  | 181 | (111-264) | 0.087 |  | 70 | (55-93) | 0.916 | **<0.001** |
| At last lab draw |  | 117 | (85-232) |  |  | 70 | (57-89) |  | **<0.001** |
| Total bilirubin | 33 |  |  |  | 32 |  |  |  |  |
| At first lab draw |  | 0.8 | (0.5-1.2) | 0.152 |  | 0.7 | (0.4-0.8) | 0.59 | 0.128 |
| At last lab draw |  | 0.5 | (0.4-0.8) |  |  | 0.5 | (0.4-0.8) |  | 0.763 |
| * comparing first and last lab draw; ^ comparing male and female values |  |  |  |  |  |  |  |  |  |

#### LV Structure

In regard to LV remodeling, LV dilation occurs in both females and males over time, however, similar to LVEF, based on mixed model coefficients and non-overlapping confidence intervals, LV dilation is more pronounced in patients who experience a HF outcome compared to those who do not. [Figure 4] [Table 3] In mixed effect modeling of z-scores that included only pediatric echocardiograms, LVEDD increased in females who experienced a HF outcome (p<0.001). Females who did not experience a HF outcome (p=0.516) and males regardless of HF outcome (p-values: HF outcome 0.704, no HF outcome 0.157) had no significant change in LVEDD based on z-scores during their pediatric years.

**Figure 4.**
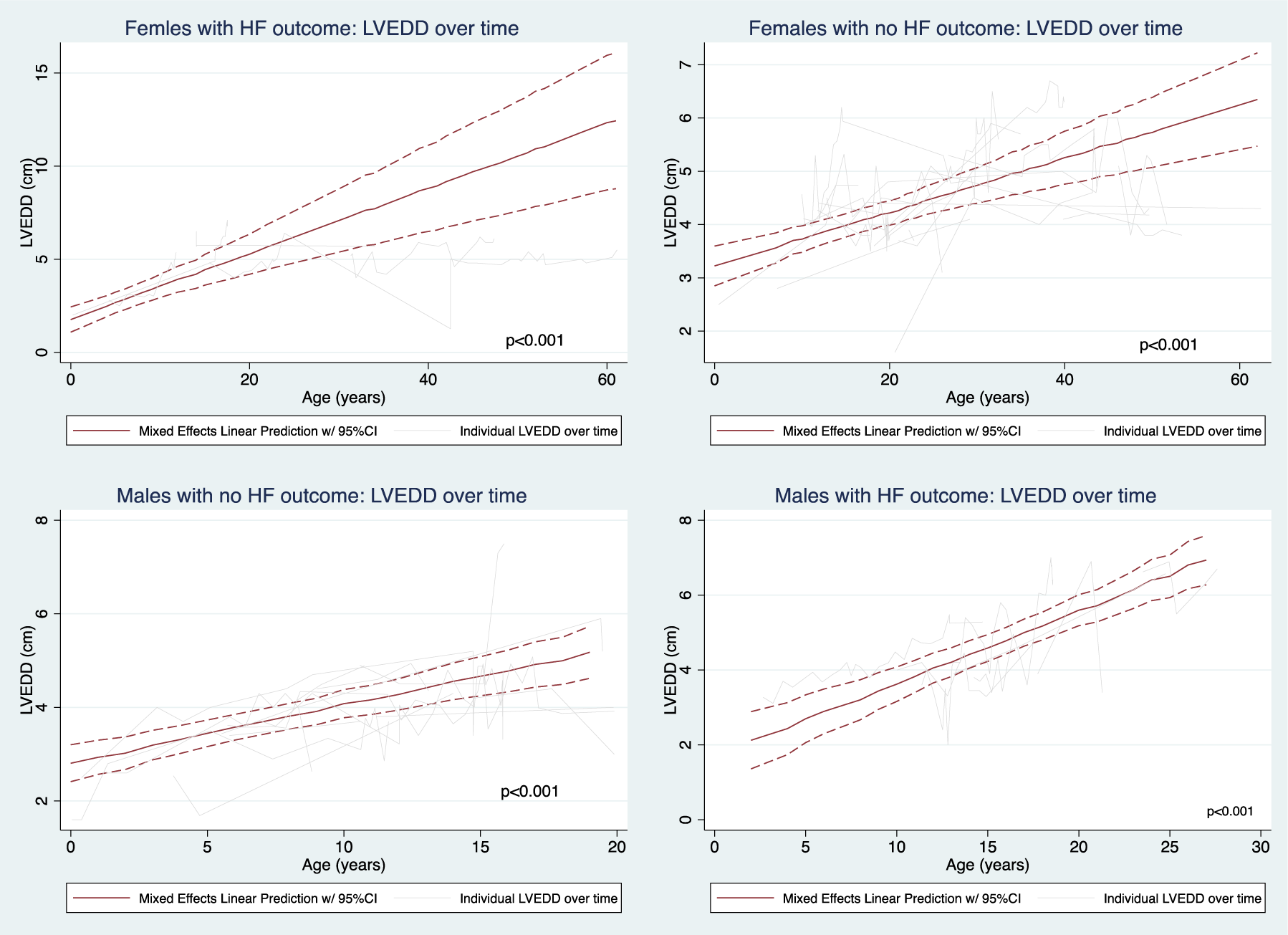
LVEDD over time by sex and HF outcome

In regards to LVH, males who experienced the HF outcome had thicker LV walls (p=0.015) on first echocardiogram, however, progressive LV hypertrophy until the time of HF outcome does not occur (p=0.734). LV hypertrophy continues in males who have not yet experienced a HF outcome (<0.001). [Table 3, Figure 5] Notably, in the mixed effect model of pediatric echocardiograms, LVH continues until the age of 18 years regardless of HF outcome (p<0.001). In females, unlike males, there is regression in LV thickness over time in both those who experience a HF outcome, as well as, those who do not. In the mixed effect model of LV wall thickness in pediatric echocardiograms only, regression in LV wall thickness was noted in females who experienced a HF outcome (p<0.001), but not in females who did not (p=0.387). [Figure 5] LV mass increases in males regardless of HF outcome and begins in childhood and continues through adulthood. In females, LV mass does not change significantly with age, likely due to progressive thinning and dilation that occurs over time in females. [Figure 6] In the pediatric z-score analysis, mixed effect model showed decreasing LV mass in females who experience a HF outcome (p=0.012).

**Figure 5.**
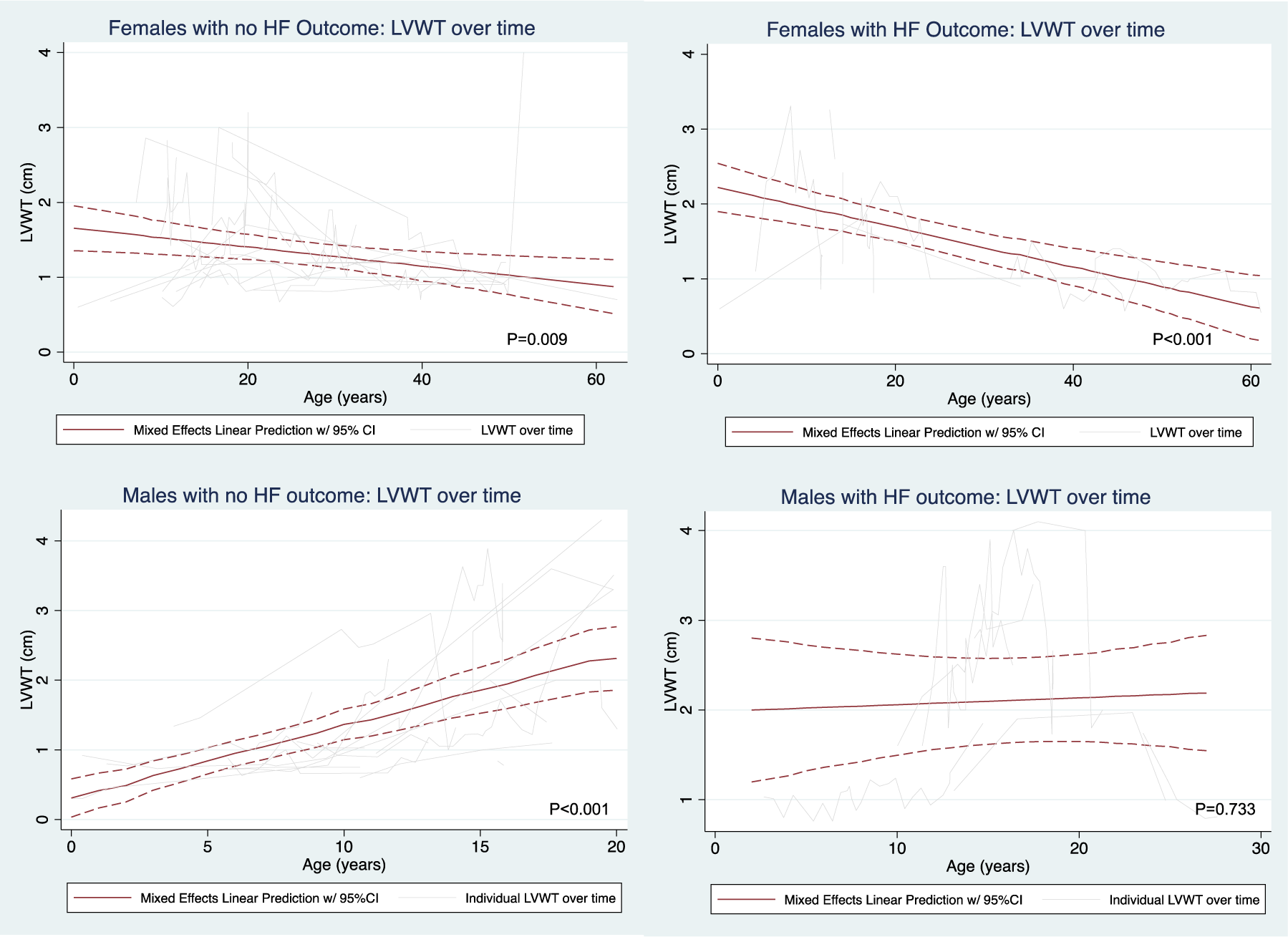
LV wall thickness measurement over time by sex and HF outcome. Thickest septal or posterior wall measurement reported.

**Figure. 6.**
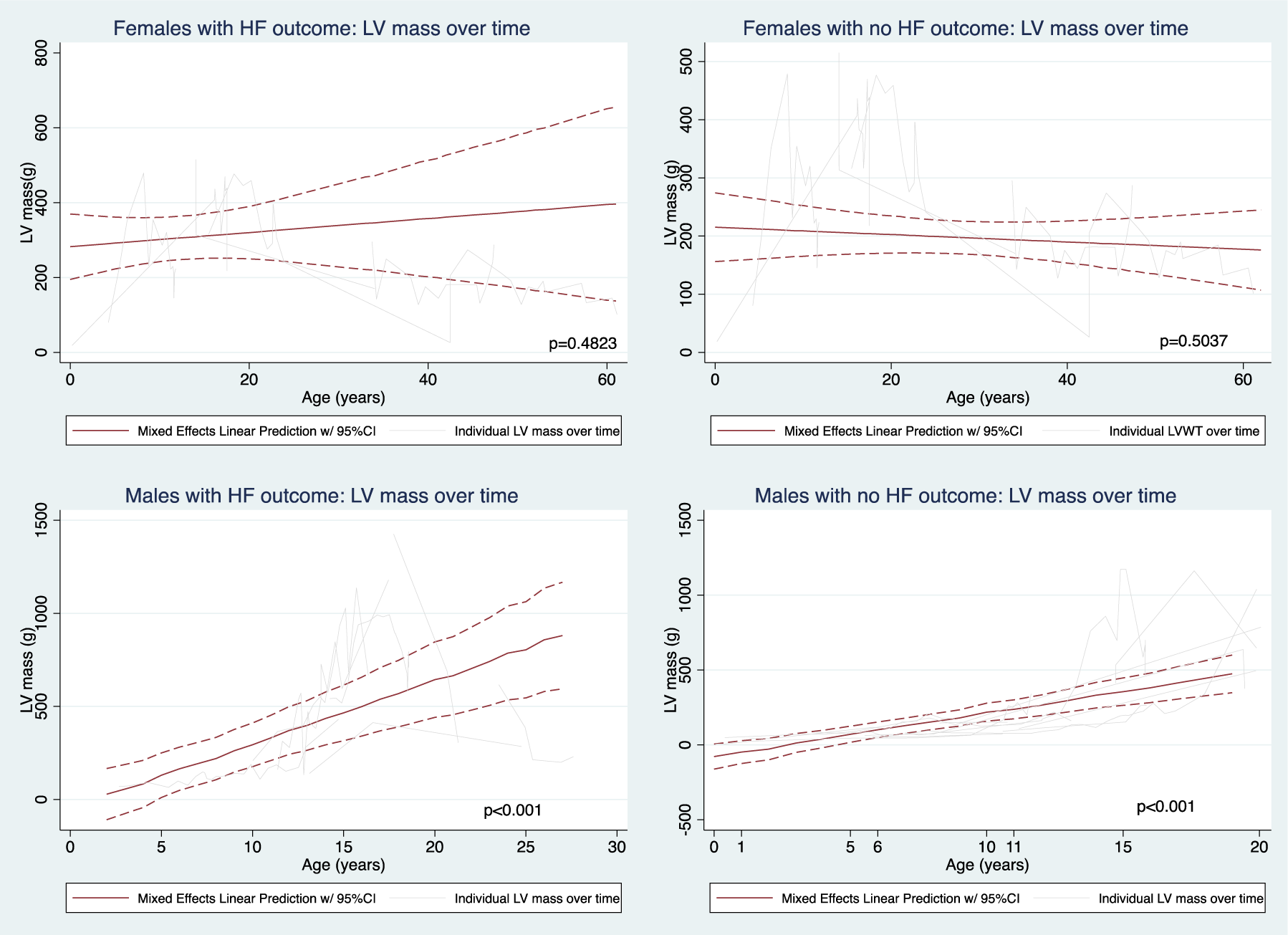
LV Mass measurement over time by sex and HF outcome

### Female stratified analyses

Given the results above, we chose to further evaluate outcomes specifically among females. In stratified analyses of females by HF outcome before 26 years old (<26 yo, n=7) and at or after 26 years old (≧26yo, n=7), age of diagnosis was significantly earlier (p=0.025) in the <26yo cohort (14 years (10 – 20 years)) compared to the ≧26yo cohort (34 years (28 – 40 years)). Regarding clinical characteristics, prevalence of CIED (86% vs 43%, p=0.094) and WPW (43% vs 0%, p=0.051) were numerically higher in the <26yo compared to ≧26yo groups. Analysis of echocardiographic parameters suggests increased LV hypertrophy in the <26yo compared to ≧26yo group, with higher numerical values at both first and last echocardiogram [Table 6]. Notably, although all females who reached a HF outcome met criteria for LV hypertrophy at some point during their life, patients in the <26yo group developed more significant hypertrophy compared to those in the ≧26yo (2.36cm (2.30 – 3.26 cm) vs 1.4cm (0.9 – 1.5 cm); p=0.020). In regards to overall longitudinal change in LVWT, in the <26yo group there was no change in LVWT as these females aged, while in the ≧26yo, there was LV thinning over time.

**Table 5.**
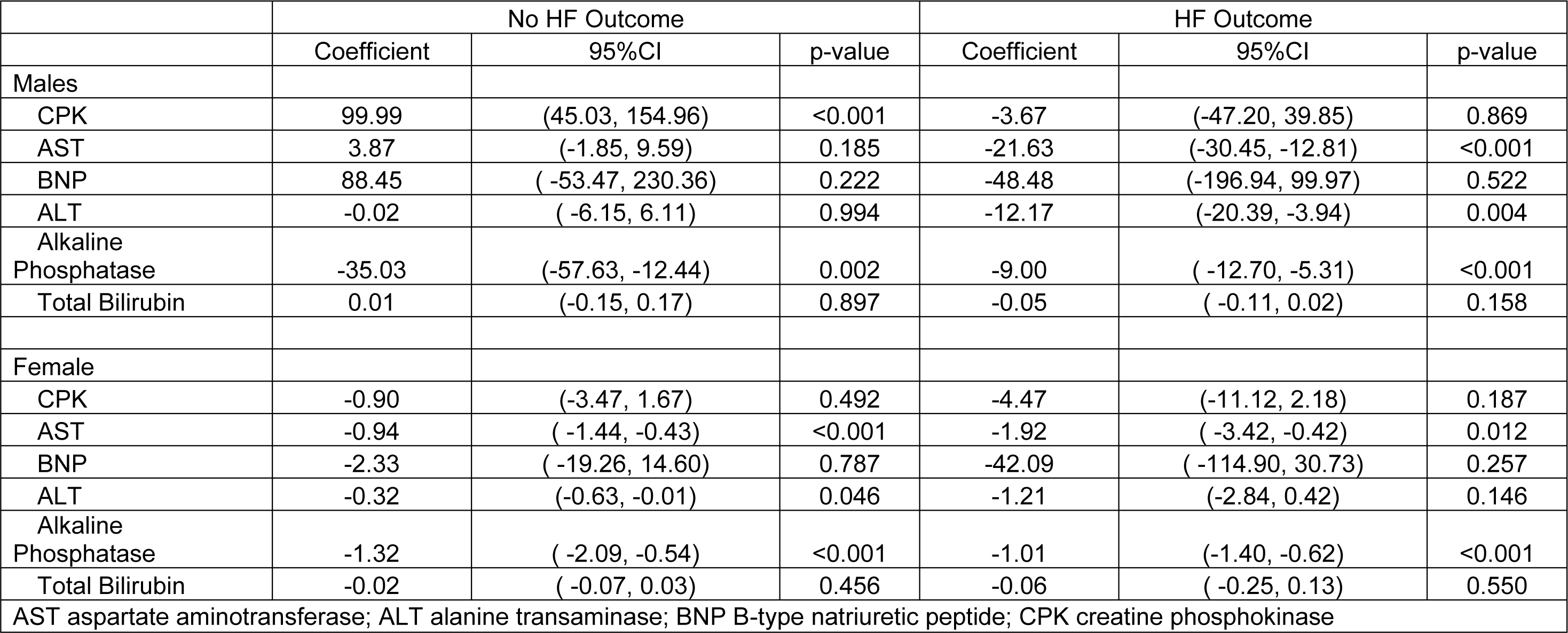
Longitudinal trend analysis of laboratory parameters over time. Coefficients derived from mixed effect regression odels represent the magnitude of change in the parameter as patient ages.

**Table 6.** Echocardiographic measurements in females who experienced a HF outcome at first and last exam stratified by ge of HF outcome. Between group comparisons for first and last measurement were completed and p-values reported.

|  | <26 years old |  | ≥26 years old |  |  |
| --- | --- | --- | --- | --- | --- |
|  | Median | IQR | Median | IQR | p-value |
| LVEF (%) |  |  |  |  |  |
| First | 77 | (52-78) | 55 | (35-57) | 0.302 |
| Last | 37 | (23-55) | 30 | (22-39) | 0.739 |
| Thickest LV wall (cm) |  |  |  |  |  |
| First | 1.4 | (1.1-2.3) | 1.0 | (0.90-1.02) | 0.121 |
| Last | 1.5 | (1.0-2.3) | 0.83 | (0.55-1.10) | 0.182 |
| LV Mass (g) |  |  |  |  |  |
| First | 318 | (80-381) | 170 | (26-296) | 0.302 |
| Last | 232 | (220-277) | 194 | (101-287) | 0.643 |
| LVEDD (cm) |  |  |  |  |  |
| First | 3.9 | (2.8-4.8) | 5.2 | (1.3-5.7) | 0.796 |
| Last | 6.1 | (5.6-6.6) | 5.8 | (5.5-6.1) | 0.643 |
| LVEF left ventricular ejection fraction; LVEDD left ventricular end diastolic dimension |  |  |  |  |  |

[Table 7] If longitudinal change in LVWT in the <26yo group was measured starting from the echocardiogram with the greatest LV hypertrophy, females in the <26yo group (mixed effects coefficient: −0.3258 (−0.434, −0.218); p<0.001) were noted to have ventricular thinning similar to the ≧26yo group prior to experiencing a HF outcome. At individual levels, some females (including those who experienced HF outcomes in the <26yo group) appear to have had progressive hypertrophy with subsequent decreases in LVWT and LVM. [Figures 5-7] LVEF decreased in both groups, however, the progression of LV dysfunction was more pronounced in the <26yo compared to the ≧26yo group. LVEDD increased in both groups, however, the rate of dilation over time was higher in the <26yo compared to the ≧26yo group. In longitudinal analysis, LV mass continued to increase in the <26yo group, while in the ≧26yo LV mass remained unchanged. [Table 7].

**Figure. 7.**
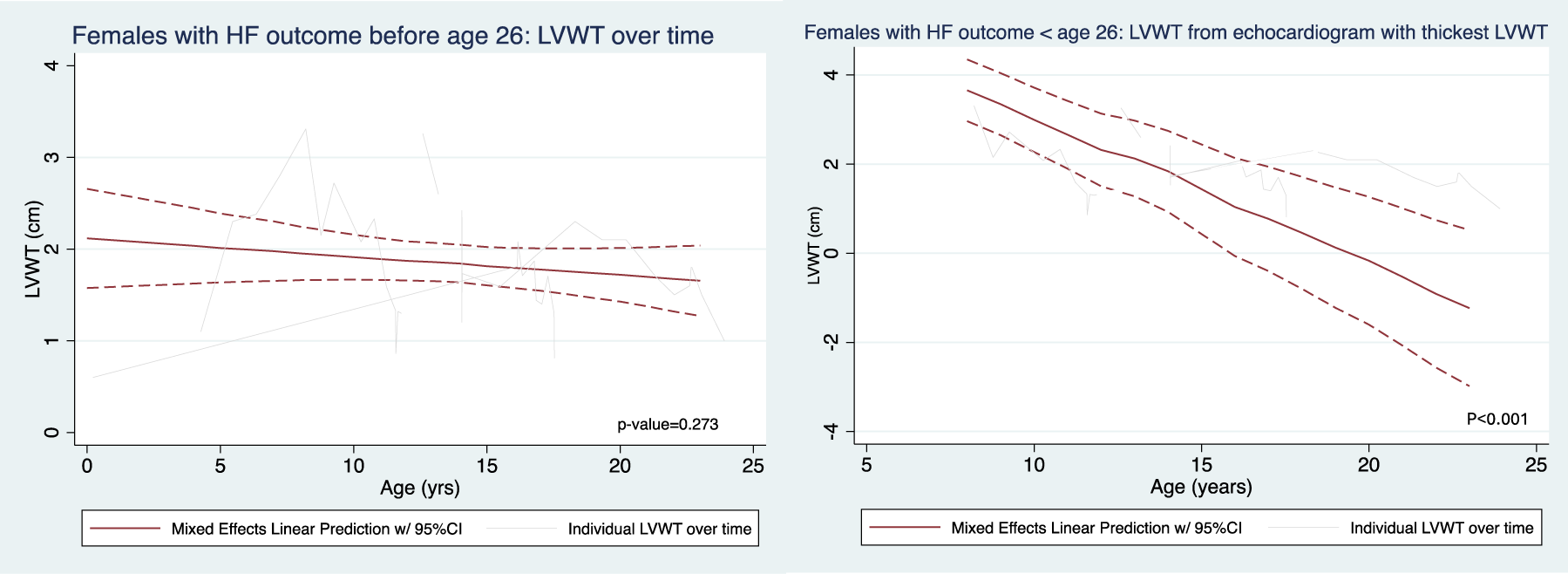
LVWT in females who experience HF outcome prior to 26 years of age at all time points and from time of thickest LVWT.

**Table 7.** Longitudinal trend analysis of echocardiographic parameters over time in female patients who experienced a HF utcome stratified by age of HF outcome. Coefficients derived from mixed effect regression models represent the agnitude of change in the parameter as patient ages.

|  | <26 years old |  |  |  |  | ≥26 years old |  |  |  |  |
| --- | --- | --- | --- | --- | --- | --- | --- | --- | --- | --- |
|  | Patients<br>(n) | Values<br>(n) | Coefficient | 95%CI | p-value | Patients<br>(n) | Values<br>(n) | Coefficient | 95%CI | p-value |
| LVEF* | 6 | 42 | -3.285 | (-4.690, -1.880) | <0.001 | 3 | 30 | -1.239 | (-1.684, -0.793) | <0.001 |
| LV wall thickness | 6 | 45 | -0.020 | (-0.055, 0.016) | 0.273 | 3 | 31 | -0.021 | (-0.033, -0.009) | <0.001 |
| LV mass* | 6 | 42 | 9.800 | (3.440, 16.160) | 0.003 | 3 | 30 | -1.913 | (-4.520, 0.695) | 0.151 |
| LVEDD* | 6 | 42 | 0.249 | (0.183, 0.315) | <0.001 | 3 | 34 | 0.068 | (0.028, 0.109) | 0.001 |
| *significant difference in coefficients by age of HF outcome based on non-overlapping 95% confidence intervals (CI) |  |  |  |  |  |  |  |  |  |  |

### Laboratory Analyses

The availability of laboratory values differed by lab test and sex. For males, availability of laboratory values ranged from 44-63% and in females 31-50%. [Table 4] None of the skeletal muscle, cardiac and liver biomarkers changed between first and last measurement. First laboratory values in patients revealed higher CPK, AST, ALT and alkaline phosphatase in males compared to females (p-value<0.001). In longitudinal analyses by mixed effect models, in males, CPK decreased in patients who did not experience a HF outcome (p<0.001) and AST and ALT decreased in patients who experienced a HF outcome (p<0.001 and p=0.004 respectively). Alkaline phosphatase decreased regardless of HF outcome in males (p<0.01). In females, AST and alkaline phosphatase decreased in all patients regardless of HF outcome (p<0.02), and ALT decreased only in females who did not experience a HF outcome (p=0.046). [Table 5].

## DISCUSSION

To date, while there are several natural history studies and meta-analyses that provide cross sectional analyses of patient characteristics and outcomes, none include longitudinal echocardiographic data that parallel disease progression.^2, 5–8, 10^ This current study analyzes the largest DD cohort to date and is the only study that provides insight into longitudinal changes in echocardiographic parameters with disease progression. By stratifying analyses by HF outcome, this study provides insight into what changes in specific echocardiographic parameters may suggest worsening disease.

### Heart Failure Outcomes

As an X-linked disease, disease penetrance in males with DD is complete, and males will express a severe disease phenotype that results in death in the absence of left ventricular assist device or cardiac transplantation by the 2^nd^ or 3^rd^ decade of life.^2, 5–8,10^ Consistent with this expected disease course, the incidence of HF outcome in our cohort was greater in males compared to females. Similar to our previously published data on DD patients who underwent transplantation, while there was a trend towards HF outcome occurring at an earlier age in males compared to females, this did not reach statistical significance.^4^ This is likely due to there being a distinct subset of females who experience a HF outcome at an earlier age in the absence of advanced heart failure therapies. In this study group, when female patients who reached HF outcome were stratified into early (less than 26 years old) and late (greater than or equal to 26 years old) HF outcome groups, 2 distinct age groups were noted (p=0.002). Specifically, median age at time of HF outcome in the early and late groups were 14 years and 39 years, respectively. Further research is needed to better characterize and identify females who are at risk of a malignant clinical course similar to males.

### Echocardiographic Changes Over Time

A corollary to characterizing longitudinal changes in LV structure over time is how this data can impact clinical decision making. Specific to our findings is utilizing changes in LVEF and structure to assist in patient management, risk stratification as well as identification of surrogate markers of disease progression and clinical outcomes. Our primary longitudinal findings are the differential changes in LV function and structure that may discriminate patients who will experience a HF outcome from those who will not.

Specifically, while LVEF drops irrespective of sex and HF outcome over time, the degradation of LVEF is more accelerated in those who experience a HF outcome. Similarly, while LV dilation occurs regardless of sex and HF outcome, which may be due to age-appropriate growth in this patient population that includes pediatric and adult data, the rate of LV dilation is also higher in those who experience a HF outcome. This may be contributed to by decreases in LVEF and stabilization of LVH in males who experience a HF outcome and also observed thinning in females regardless of HF outcome. These findings suggest that LVEF and LVEDD may be used to predict HF outcome and can be used by clinicians to escalate surveillance intervals and risk stratify DD patients for advanced therapies. Notably, progressive disease in hypertrophic cardiomyopathy is also marked by increasing LVEDD and also by decreased LVEF.^11^

A major difference in phenotypes between sexes is that in males who have not yet experienced HF outcome, LV hypertrophy continues. This suggests that eventual LVH stabilization may be a marker for end-stage disease in males particularly when paired with progressive LV dysfunction and dilation. The pattern of LV remodeling in this cohort of females was different, with LV thinning occurring over time regardless of HF outcome. Consistent with the aforementioned differences in LV remodeling between males and females, LV mass increases over time in males, while no change was found in females. Of note, the analyses stratifying females by age of HF outcome suggests that there may be a female subgroup who are phenotypically similar to males, experiencing end-stage heart failure or death earlier in life. Females who experience a HF outcome earlier in life have more severe LV hypertrophy compared to females who experience a HF outcome later in life. Additionally, like males, their LV mass increases significantly over time. These finding highlight the importance of additional studies elucidating varying Danon disease phenotypes and mechanisms impacting disease expression in females.

Analyses of normalized pediatric echocardiographic parameters provides some insight into disease progression as trends in LV hypertrophy and increasing LVEDD through adulthood were not seen during the pediatric years, suggesting that several of the variables that are seen in patients with HF outcome likely occur later in the disease process. Prior case report studies utilizing cardiac MRI for tissue characterization, have shown that increased late gadolinium enhancement and mass have been associated with a composite endpoint that includes death, heart transplantation and ICD implantation for secondary prevention.^12–14^ Furthermore, a study evaluating longitudinal strain in Danon disease patients found that strain was similarly associated with death and advanced heart failure requiring either transplantation or ventricular assist device.^15^ Additionally longitudinal studies to assess how progressive fibrosis may be contributing to these sex specific changes with disease progression are needed.

### Laboratory Changes Over Time

Interpretation of laboratory values is limited by sample size, with majority of laboratory values presented having ∼50% completion, and the selection bias in those who ended up getting repeat laboratory studies completed. To this end, interpretation of longitudinal trends is challenging and instead will highlight the cross-sectional analysis that showed that CPK, AST and ALT were all markedly elevated in males compared to females.

Notably, total bilirubin was neither elevated nor different between sexes and alkaline phosphatase, while higher in males compared to females, was only mildly elevated. These patterns suggest that the CPK, AST and ALT elevations are likely explained by increased severity of skeletal myopathy in males compared to females.

### Clinical Trial Design

Additionally, although cardiac transplantation remains the only therapeutic option with a survival benefit, a Phase I gene therapy trial in DD has completed and a Phase II AAV9 gene therapy trial is pending enrollment.^16^ Major barriers to designing a DD comparative effectiveness trial with a traditional control population include low disease prevalence, incidence of outcomes and treatment effect sizes. To mitigate the effect of these barriers, the FDA has encouraged: 1) non-traditional trial design, including the use of historical controls and 2) pathways for accelerated drug approval which include using surrogate endpoints that predict traditional clinical outcomes associated with improved patient functional status, quality of life and survival.^17,18^ To this end, natural history studies are an important adjunct to understanding disease course and developing therapeutics that will impact disease progression and outcomes. The results from this study suggest that potential surrogate endpoints for therapeutic trials could include LVEF, LV mass, LVEDD and LV wall thickness in males, and LVEF, LVEDD and LV wall thickness in females. Additionally, because LV hypertrophy was a prominent feature in those experiencing a HF outcome, particularly in males and also females who experienced a HF outcome earlier in life, trial design including hypertrophy as an entry criteria can be considered.

### Limitations

While this study reports outcomes from the largest cohort of Danon Disease patients, the sample size remains small and limits generalizability. Patient registries often suffer from variability in data entry and due to the retrospective nature of the study and also standard of care data collection that spanned enrollment over several decades, data collection intervals and parameters were not consistent. Notably, echocardiography was obtained in >80 percent of the patient population, however, laboratory values were only collected in ∼50%, with CPK and BNP only collected in ∼30% of female patients. All patients included within the study had outcomes data collected though. Even with these limitations, this remains the largest cohort with longitudinal echocardiographic and laboratory data.

## Data Availability

All data is available from International Danon Disease Registry.

---

| **Abbreviations Table** |  |
| --- | --- |
| Danon disease | DD |
| left ventricular | LV |
| lysosome-associated membrane protein-2 | LAMP-2 |
| ejection fraction | EF |
| left ventricular end diastolic diameter | LVEDD |
| left ventricular septal wall thickness | LVSW |
| left ventricular posterior wall thickness | LVPW |
| left ventricular hypertrophy | LVH |
| left ventricular assist device | LVAD |
| heart failure | HF |
| transplantation | TXP |
| cardiovascular implantable electronic device | CIED |
| atrial fibrillation | AF |

